# A Comparative Analysis of In-Hospital Outcomes and Care Cost Between Surgical and Transcatheter Valve Replacement for Aortic Stenosis: Insights From the U.S Nationwide Inpatient Sample Database

**DOI:** 10.1101/2023.08.25.23294647

**Authors:** Fidelis E. Uwumiro, Michael Bojerenu, Charles T. Ogbodo, Victory O. Okpujie, Cherechi O. Nwabueze, Emmanuel O. Otabor, Muhammed L. Shielu, Chuka G. Nwume, Omolade J. Oshodi, Hillary Alemenzohu, Olawale O. Abesin

## Abstract

**Background/objectives:** Transcatheter aortic valve replacement (TAVR) has emerged as a preferred alternative to surgical aortic valve replacement (SAVR) for symptomatic aortic stenosis. This study aimed to compare the clinical outcomes and care costs of TAVR and SAVR to medical management using five years of inpatient data.

**Methods:** Adult hospitalizations with a principal diagnosis of aortic stenosis were analyzed from the Nationwide Inpatient Sample database (2016-2020). Diagnosis and procedure variables, as well as confounders and comorbidities, were identified using the International Classification of Diseases (ICD-10) codes. Multivariable regression models were utilized to assess mortality odds, length of stay (LOS), periprocedural complications, and care costs.

**Results:** Among the 364,515 admissions for aortic stenosis analyzed, the mean age was 76 ± 0.5 years, with a majority of male patients (57.8%) and White Americans comprising 85.5% of the population. SAVR was performed in 29.3% of cases, and TAVR in 50.8%. TAVR demonstrated significantly lower in-hospital mortality compared to SAVR (aOR: 0.463; 95% CI: 0.366-0.587; P < 0.001), whereas SAVR did not show a significant difference (aOR: 0.786; 95% CI: 0.601-1.029; P = 0.079). TAVR also resulted in a significantly shorter mean LOS compared to SAVR (adjusted mean LOS: 2.37; 95% CI: 2.12-2.63; P < 0.001 vs. 6.25; 95% CI: 6.00-6.50; P < 0.001). While TAVR patients had a lower likelihood of complications, they incurred higher hospital costs.

**Conclusion:** TAVR demonstrated significantly lower odds of in-hospital mortality and shorter length of stay compared to medical management or SAVR. However, TAVR patients incurred higher hospital costs despite a lower likelihood of complications.

## INTRODUCTION

Both surgical aortic valve replacement (SAVR) and transcatheter aortic valve replacement (TAVR) remain viable alternatives for treating symptomatic aortic stenosis (AS). SAVR, the traditional surgical approach, involves a sternotomy or minimally invasive thoracotomy to replace the diseased valve with a prosthetic valve. On the other hand, TAVR represents a less invasive percutaneous procedure in which an expandable transcatheter valve is delivered to the site of aortic stenosis through a femoral or transapical access, thereby avoiding open-heart surgery.

In recent years, extensive research has increasingly demonstrated the efficacy of TAVR as a favorable alternative to medical therapy, while also exhibiting comparable effectiveness to SAVR in the treatment of severe AS patients with intermediate and high surgical risk.^1–3^ Notably, TAVR has exhibited comparable mortality and stroke rates in high-risk surgical patients.^4^ These favorable outcomes have resulted in the widespread adoption of TAVR as the primary method for aortic valve replacement in the United States.^5^ However, given that a significant proportion of patients with severe symptomatic AS fall into the low-risk category (approximately 40% of the population),^6^ and considering the substantial cost associated with TAVR technology, it becomes crucial to determine the optimal treatment approach for each specific patient scenario. This determination should consider both clinical outcomes and economic burden.

While numerous studies have examined the clinical outcomes and procedural efficacy of SAVR and TAVR individually, there remains a need for a comprehensive comparative analysis that evaluates both clinical outcomes and care costs associated with these two treatment modalities. Such a study would provide valuable insights to guide clinical decision-making, inform healthcare resource allocation, and improve patient-centered care.

Therefore, the objective of this study is to perform a comparative analysis of outcomes and care costs among patients admitted for AS who receive SAVR or TAVR. By analyzing a comprehensive dataset that incorporates patient and hospital-level factors, along with cost information, this study aims to provide insights into the clinical outcomes, safety, and economic implications of these treatment modalities. Specifically, we intend to assess and compare the likelihood of mortality, periprocedural complications, duration of hospitalization, and associated hospital charges for both approaches. We hypothesize that TAVR will demonstrate superior outcomes to SAVR.

## METHODS

### Data source

The study drew upon data obtained from the Nationwide Inpatient Sample (NIS) database, spanning the years 2016 to 2020. The NIS represents an expansive collection of inpatient stays across the United States, standing as the largest publicly accessible repository. It contains discharge data from a 20% stratified sample of community hospitals and is a part of the Healthcare Quality and Utilization Project (HCUP), sponsored by the Agency for Healthcare Research and Quality (AHRQ).^7^ Within this rich dataset, comprehensive information is available, including both the primary and up to 25 secondary diagnoses, patient demographics as well as procedures, with a capacity for recording up to 15 primary and secondary procedures. Additionally, the database provides valuable insights into key hospital characteristics such as ownership, bed size, teaching status, urban/rural location, and region. Additionally, it affords access to healthcare resource utilization data, such as length of hospital stay (LOS), total hospitalization charges, and patient discharge disposition. The dataset encompasses all hospital stays, extending its coverage to encompass even Medicare-advantage patients, who constitute up to 30% of the entire Medicare beneficiary population.^8^ Hospitalizations within the NIS database are recorded using the International Classification of Diseases, Clinical Modification and Procedure coding system (ICD-10-CM/PCS). The large sample size and nationwide scope of the NIS present a unique opportunity to investigate distinctive medical procedures, treatments, and rare patient groups. It also allows researchers to test findings from single-center studies or smaller patient populations on a more diverse and representative patient population.

### Ethical consideration

The use of the NIS is regulated by the U.S. AHRQ. The current design of the NIS meticulously adheres to the stipulations set forth in the Healthcare Insurance Portability and Accountability Act (HIPAA) of 1996 and the Omnibus Final Rule of 2013. Commencing in 2012, the AHRQ has implemented the exclusion of 16 direct identifiers from the NIS dataset, thereby ensuring the preservation of patient and hospital information by upholding the highest privacy and safety standards. Consequently, the NIS is classified as a limited data set, thereby exempting it from the requirement of Institutional Review Board (IRB) approval.^9,10^

### Patient population and study design

We identified all adult hospitalizations for individuals with a principal diagnosis of symptomatic AS utilizing the relevant ICD-10-CM/PCS codes (I35.0, I06.0). To ensure the reliability of our findings, we excluded patients under the age of 18, patients with a prior history of heart surgery, or presence of prosthetic heart valves (ICD-10 codes: Z95.2, Z48.812, Z92.4, Z87.74, and Z94.1) and admissions with incomplete or missing data from our study cohort. The total cohort was further divided based on the procedural approach for aortic valve replacement into two distinct subgroups: SAVR and TAVR. These subgroups were identified using specific ICD-10 codes: 02RF07Z, 02RF08Z, 02RF0JZ, and 02RF0KZ for SAVR, and 02RF37H, 02RF37Z, 02RF38H, 02RF38Z, 02RF3JH, 02RF3JZ, 02RF3KH, and 02RF3KZ for TAVR. Admissions without evidence of valve replacement, representing cases managed solely through medical approaches, were included as controls in our study.

**Figure 1:**
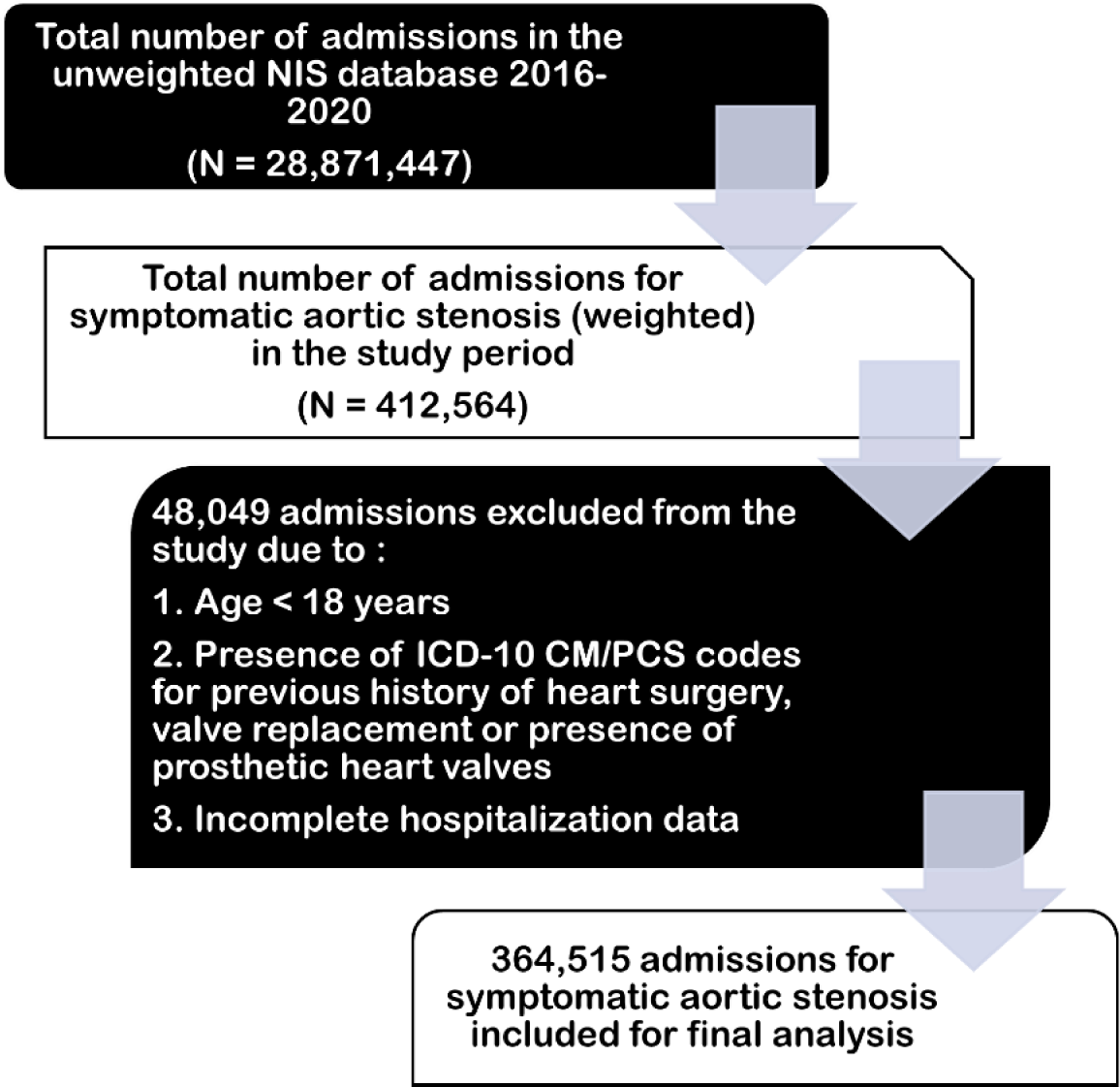
Inclusion and exclusion criteria for the study. Shows stepwise selection criteria for the final study cohort for data analysis ICD-10 CM/PCS, International Classification of Diseases, Clinical Modification/Procedure Coding System; NIS, nationwide inpatient sample

### Outcome measures and study variables

The main objective of our study was to assess the likelihood of inpatient mortality among the distinct subgroups and control subjects. In the NIS database, mortality is documented as a binary variable (designated as “DIED”). Additionally, we explored several secondary outcomes to gain further insights, including the average length of stay (LOS), mean total hospital charges (TOTCHG), and periprocedural complications. The LOS and TOTCHG variables are pre-defined numerical indicators readily available within the NIS database. To identify the most prevalent complications following aortic valve replacement, we referred to existing literature.^11–14^ Subsequently, we included these complications in our analysis exclusively if they possessed specific ICD-10-CM/PCS codes that had been validated in prior research. Other relevant variables included sociodemographic variables such as age, gender, race, median annual income quartiles specific to the patient’s ZIP code, as well as hospital-related factors including region, bed size, and teaching status. These variables were already pre-established and readily accessible within the NIS database.

### Statistical analysis

All analyses were conducted using Stata, version 17.0BE (StataCorp LLC, College Station, TX, USA), adhering to HCUP recommendations for calculating national estimates. To ensure accurate nationwide estimations, analyses were performed on a weighted sample representing over 35 million hospitalizations annually.

Unadjusted odds ratios (ORs) for the primary outcome were initially calculated through univariable logistic regression analyses, considering all variables listed in Table 1. Subsequently, variables with p-values below 0.1 were selected for inclusion in the final multivariable logistic and regression models. This selection criterion, taking into account the large sample size, aimed to avoid the inclusion of marginally related factors that do not significantly impact the outcome of interest. To identify established confounders related to the primary and secondary outcomes, a comprehensive review of existing literature was conducted. These confounders included age ≥ 80, early surgery (aortic valve replacement performed within 3 days of hospitalization), frail phenotype (defined using the Johns Hopkins Adjusted Clinical Groups frailty clusters),^15,16^ obesity, history of ischemic heart disease, diabetes mellitus, the incidence of any complication or multiple procedures in the index admission, and prolonged hospitalization (defined as hospital length of stay in the top decile of all admissions in the study cohort).

**Table 1.**
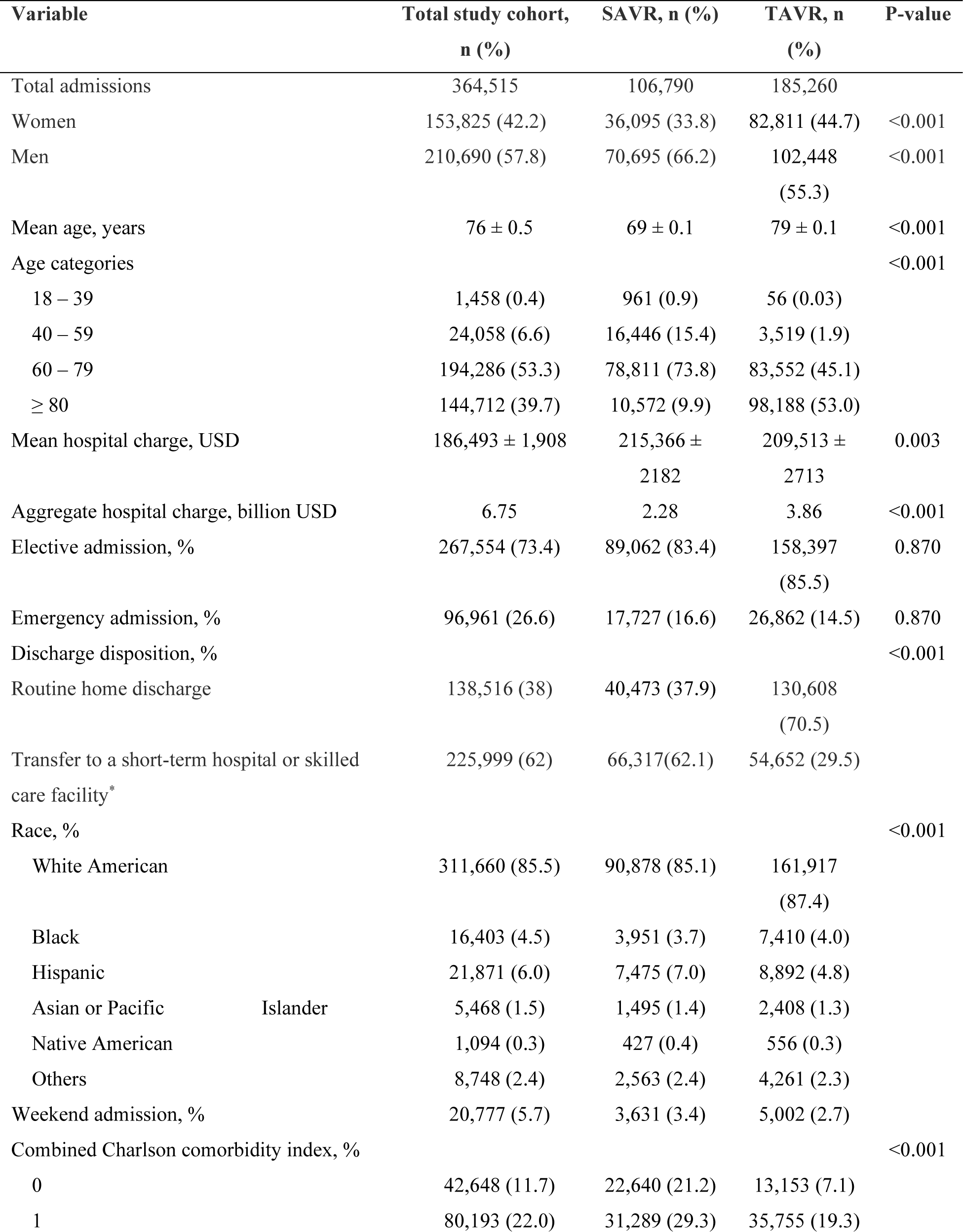
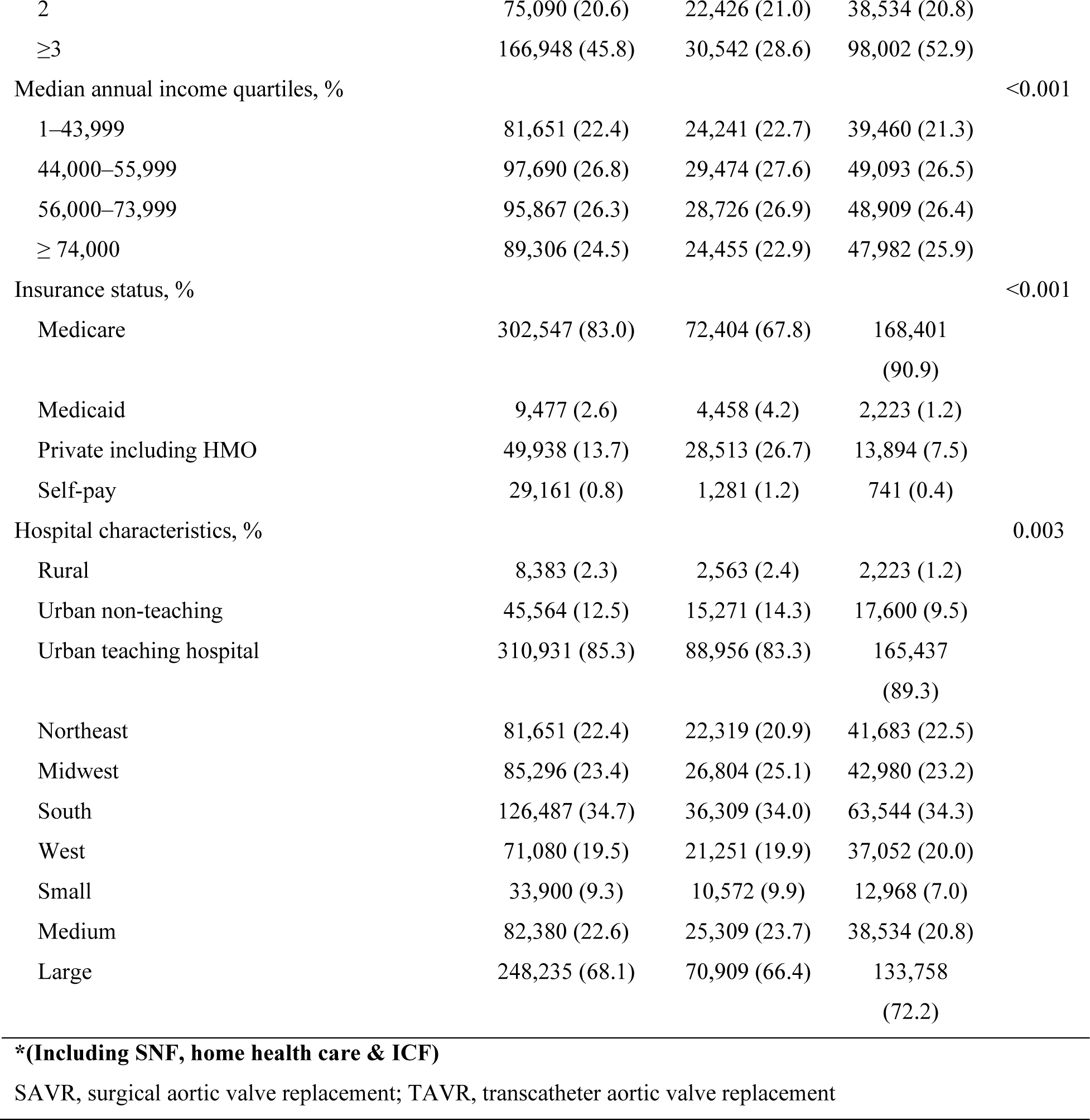
Baseline sociodemographic composition and resource utilization of aortic stenosis hospitalizations.

The burden of co-morbidity was assessed using Sundararajan’s adaptation of the modified Deyo’s Charlson Comorbidity Index (CCI), categorized into four groups reflecting escalating mortality risk. A CCI score exceeding 3 corresponds to an approximate 25% 10-year mortality rate, while scores of 2 or 1 correspond to 10% and 4% 10-year mortality rates, respectively.^17,18^

We compared baseline sociodemographic characteristics between TAVR and SAVR cohorts using Pearson’s chi-square tests. Proportions between nominal variables were compared using chi-square tests, while Student’s t-test was utilized for continuous variables. The significance level for all the multivariable regression analyses was set at p-values below 0.05. Categorical variables were reported as proportions, and continuous variables were reported as mean with standard deviation. The regression outcomes were reported as adjusted odds ratios (aORs) or β coefficients (adjusted mean difference) with 95% confidence intervals (CIs).

### Data availability statement

In compliance with the copyright restrictions set by the AHRQ pertaining to the distribution of HCUP databases, the database utilized in this research will not be made publicly available by the authors. However, all NIS datasets are publicly available through the authorized HCUP central distributor upon request at https://hcup-us.ahrq.gov/tech_assist/centdist.jsp.

## RESULTS

### Sociodemographic characteristics of the study population

During the study period, a total of 364,515 adult admissions were recorded for aortic stenosis. Among these admissions, SAVR was performed in 29.3% (106,790) of cases, while TAVR was utilized in 50.8% (185,260) of cases. The remaining 19.9% of admissions within the study cohort were managed solely through medical treatments. The mean age of the study population was 76 ± 0.5 years, with the SAVR and TAVR subgroups having mean ages of 69 ± 0.1 years and 79 ± 0.1 years, respectively. The majority of the study cohort consisted of men, representing 66.2% of the SAVR subgroup, 55.3% of the TAVR subgroup, and 57.8% of the entire study cohort. Additionally, approximately 93% of all admissions were individuals aged 60 years or older. Notably, the majority of patients undergoing SAVR fell within the age range of 60-79 years (73.8%), whereas the majority of TAVR recipients were aged 80 years or older (53%).

Regarding the racial composition of the study population, the majority were White Americans, accounting for 85.5% of the cohort. Furthermore, most admissions occurred electively (73.4%) on weekdays and were concentrated in large urban teaching hospitals located in the southern and midwestern states (Table 1). A significant proportion of the total study cohort (66.4%) exhibited a combined Charlson comorbidity index (CCI) score of 2 or higher. Specifically, 52.9% of patients who received TAVRs had a CCI score of ≥ 3. In terms of insurance coverage, Medicare was the most prevalent form of insurance across all subgroups, accounting for 83%, 67.8%, and 90.9% of admissions in the total, SAVR, and TAVR groups, respectively. Private insurance (including Health Maintenance Organizations) was the second most common payer across all study groups.

### Inpatient mortality

A total of 7,335 mortalities, constituting 2% of the study population, were documented in this study. Among these fatalities, 28.6% (2095) occurred in patients who underwent SAVR, while 29.9% (2190) were observed in individuals who received TAVR. The majority of the deceased were males (51.9%) within the age range of 60 to 79 years (43.1%) or 80 years and above (46.9%). Patients with a CCI score of 3 or more accounted for the highest proportion of mortalities (63.7%). Furthermore, a significant number of mortalities were reported among individuals insured by Medicare (84.1%) and admitted to large hospitals (68.1%), predominantly situated in the Southern or Western regions. Regarding temporal trends, a greater percentage of mortalities occurred on weekdays (89.2%) compared to weekends (10.8%), with the majority transpiring in urban teaching hospitals (82.7%).

The crude odds ratio for mortality among SAVR recipients was 0.96 (95% CI: 0.86 – 1.08; P = 0.546), while for TAVR recipients it was 0.40 (95% CI: 0.36 – 0.45; P < 0.001). After adjusting for relevant patient and hospital-level factors, TAVR demonstrated a statistically significant reduction in the odds of in-hospital mortality (aOR: 0.463; 95% CI: 0.366 – 0.587; P < 0.001). Conversely, SAVR did not exhibit a significant difference in the odds of mortality in this study (aOR: 0.786; 95% CI: 0.601 – 1.029; P = 0.079).

Other independent predictors of in-hospital mortality included female sex, advanced age, higher Charlson comorbidity index, and post-operative complications such as myocardial infarction, cardiac tamponade, critical care admission, acute respiratory distress syndrome, and acute respiratory failure, all of which increased the likelihood of mortality. Interestingly, obesity, diabetes mellitus, a median annual income quartile of $74,000 or higher, and performing aortic valve replacement within 3 days of admission were associated with a reduced likelihood of in-hospital mortality (Table 2).

**Table 2.** Odds of in-hospital mortality on multivariable logistic regression analysis.

| Predictors of mortality | aOR | Standard Error | P-value* | [95% Conf. interval] |  |
| --- | --- | --- | --- | --- | --- |
|  |  |  |  | Lower limit | Upper limit |
| TAVR | 0.463 | 0.056 | <0.001 | 0.366 | 0.587 |
| SAVR | 0.786 | 0.108 | 0.079 | 0.601 | 1.029 |
| Female sex | 1.284 | 0.082 | <0.001 | 1.133 | 1.454 |
| Age | 1.035 | 0.005 | <0.001 | 1.026 | 1.044 |
| Myocardial infarction | 1.585 | 0.208 | <0.001 | 1.225 | 2.050 |
| Cardiac tamponade | 6.934 | 1.301 | <0.001 | 4.8 | 10.016 |
| Infective endocarditis | 1.829 | 0.919 | 0.229 | 0.683 | 4.897 |
| Operative wound dehiscence | 1.404 | 0.833 | 0.567 | 0.439 | 4.489 |
| Critical care admission | 10.892 | 0.953 | <0.001 | 9.176 | 12.929 |
| Ischemic heart disease | 1.159 | 0.096 | 0.075 | 0.985 | 1.364 |
| Obesity | 0.690 | 0.07 | <0.001 | 0.566 | 0.841 |
| Diabetes mellitus | 0.680 | 0.067 | <0.001 | 0.561 | 0.824 |
| Acute respiratory distress syndrome | 9.458 | 3.919 | <0.001 | 4.198 | 21.308 |
| Acute respiratory failure | 3.803 | 0.329 | <0.001 | 3.211 | 4.505 |
| Weekend admission | 0.973 | 0.101 | 0.795 | 0.794 | 1.194 |
| Race |  |  |  |  |  |
| Blacks | 0.801 | 0.118 | 0.134 | 0.600 | 1.07 |
| Hispanics | 0.93 | 0.12 | 0.571 | 0.722 | 1.197 |
| Asian/Pacific Islanders | 0.892 | 0.223 | 0.648 | 0.546 | 1.457 |
| Native Americans | 0.283 | 0.296 | 0.227 | 0.036 | 2.198 |
| Other races | 1.021 | 0.188 | 0.911 | 0.712 | 1.464 |
| Median annual income quartile in the patients' ZIP code (\$1 – \$43,999 as reference) | | | | | |
| \$44,000 – \$55,999 | 0.899 | 0.077 | 0.216 | 0.759 | 1.064 |
| \$56,000 – \$73,999 | 0.945 | 0.086 | 0.533 | 0.791 | 1.129 |
| ≥ \$74,000 | 0.749 | 0.073 | 0.003 | 0.618 | 0.907 |
| Higher Charlson comorbidity index | 1.124 | 0.017 | <0.001 | 1.091 | 1.157 |
| Aortic replacement is done within 3 days of admission | 0.648 | 0.074 | <0.001 | 0.517 | 0.811 |
| Insurance status (Medicare as reference) |  |  |  |  |  |
| Medicaid | 0.985 | 0.198 | 0.941 | 0.664 | 1.461 |
| Private insurance including HMO | 1.088 | 0.122 | 0.455 | 0.872 | 1.356 |
| Self-pay | 1.327 | 0.463 | 0.417 | 0.67 | 2.628 |
| Hospital region (Northeast as reference) |  |  |  |  |  |
| Midwest | 1.066 | 0.11 | 0.536 | 0.871 | 1.304 |
| South | 1.219 | 0.116 | 0.037 | 1.012 | 1.469 |
| West | 1.047 | 0.112 | 0.667 | 0.849 | 1.292 |
| Hospital bed size (small hospitals as reference) |  |  |  |  |  |
| Medium | 0.792 | 0.095 | 0.053 | 0.626 | 1.003 |
| Large | 0.997 | 0.104 | 0.977 | 0.812 | 1.224 |
| Admission lasting longer than 18 days† | 0.862 | 0.093 | 0.171 | 0.697 | 1.066 |
| **TAVR, transcatheter aortic valve replacement; SAVR, surgical aortic valve replacement; HMO, health maintenance organization; aOR, adjusted odds ratio** |  |  |  |  |  |
| † Prolonged admission, defined as admission length of stay in the top decile of all admissions in the study cohort |  |  |  |  |  |

### Length of hospital stay

The average LOS in the overall study cohort was determined to be 5.1 ± 0.1 days. Patients who underwent surgical aortic valve replacement (SAVR) had a longer average LOS of 8.1 days, while those who received transcatheter aortic valve replacement (TAVR) had a shorter average LOS of 3.5 days. In comparison to patients managed solely with medical interventions, both SAVR and TAVR patients exhibited a longer average LOS (5.2 vs. 4.8 days). Notably, patients who underwent SAVR or TAVR within 3 days of hospitalization demonstrated a shorter mean LOS when compared to those who underwent the procedures at a later time (4.4 vs. 6.9 days).

After adjustments for relevant confounding factors using binomial regression analysis, TAVR was found to be associated with a significantly shorter mean LOS compared to SAVR (adjusted mean LOS: 2.37; 95% CI: 2.12 – 2.63; P < 0.001 vs. 6.25; 95% CI: 6.00 – 6.50; P < 0.001) (Table 3). Both SAVR and TAVR procedures were linked to significantly increased mean LOS when compared to medical management. Furthermore, the occurrence of complications such as cardiac tamponade, post-procedure wound dehiscence, admission to the critical care unit, acute respiratory distress syndrome, and acute respiratory failure significantly prolonged the duration of hospitalization (Table 3).

**Table 3.** Predictors of hospital length of stay on multivariable linear regression analysis.

| Covariates | Coefficient* | Standard Error | P-value | 95% Confidence Interval |  |
| --- | --- | --- | --- | --- | --- |
|  |  |  |  | Lower limit | Upper limit |
| TAVR | 2.372 | 0.130 | <0.001 | 2.118 | 2.626 |
| SAVR | 6.255 | 0.128 | <0.001 | 6.004 | 6.505 |
| Female sex | 0.194 | 0.025 | <0.001 | 0.145 | 0.243 |
| Age | 0.013 | 0.002 | <0.001 | 0.009 | 0.016 |
| Myocardial infarction | 0.138 | 0.104 | 0.184 | -0.066 | 0.342 |
| Cardiac tamponade | 2.512 | 0.470 | <0.001 | 1.591 | 3.433 |
| Infective endocarditis | 0.728 | 0.694 | 0.294 | -0.632 | 2.088 |
| Operative wound dehiscence | 6.150 | 1.476 | <0.001 | 3.257 | 9.043 |
| Critical care admission | 1.820 | 0.113 | <0.001 | 1.599 | 2.041 |
| Diabetes mellitus | -0.195 | 0.029 | <0.001 | -0.251 | -0.138 |
| Ischemic heart disease | -0.125 | 0.037 | 0.001 | -0.197 | -0.053 |
| Obesity | -0.021 | 0.039 | 0.600 | -0.097 | 0.056 |
| Acute respiratory distress syndrome | 4.557 | 1.586 | 0.004 | 1.449 | 7.665 |
| Acute respiratory failure | 1.485 | 0.101 | <0.001 | 1.287 | 1.683 |
| Weekend admission | 0.701 | 0.077 | <0.001 | 0.551 | 0.851 |
| Race |  |  |  |  |  |
| Blacks | 0.403 | 0.076 | <0.001 | 0.255 | 0.552 |
| Hispanics | 0.309 | 0.066 | <0.001 | 0.18 | 0.438 |
| Asian/Pacific Islanders | 0.043 | 0.107 | 0.688 | -0.166 | 0.252 |
| Native Americans | -0.236 | 0.250 | 0.346 | -0.727 | 0.255 |
| Other races | 0.231 | 0.085 | 0.007 | 0.064 | 0.398 |
| Median annual income quartile in the patients' ZIP code (\$1 – \$43,999 as reference) | | | | | |
| \$44,000 – \$55,999 | -0.051 | 0.039 | 0.187 | -0.128 | 0.025 |
| \$56,000 – \$73,999 | 0.028 | 0.040 | 0.483 | -0.051 | 0.108 |
| ≥ \$74,000 | 0.043 | 0.043 | 0.327 | -0.043 | 0.128 |
| Higher Charlson comorbidity index | 0.210 | 0.008 | <0.001 | 0.195 | 0.225 |
| Aortic replacement is done within 3 days of admission | -3.638 | 0.129 | <0.001 | -3.892 | -3.385 |
| Insurance status (Medicare as reference) |  |  |  |  |  |
| Medicaid | 0.237 | 0.109 | 0.03 | 0.023 | 0.451 |
| Private insurance including HMO | -0.153 | 0.045 | 0.001 | -0.241 | -0.066 |
| Self-pay | 0.007 | 0.144 | 0.96 | -0.274 | 0.289 |
| Hospital region (Northeast as reference) |  |  |  |  |  |
| Midwest | -0.154 | 0.060 | 0.01 | -0.271 | -0.036 |
| South | -0.211 | 0.053 | <0.001 | -0.315 | -0.107 |
| West | -0.360 | 0.056 | <0.001 | -0.47 | -0.25 |
| Hospital bed size (small hospitals as reference) |  |  |  |  |  |
| Medium | 0.383 | 0.068 | <0.001 | 0.249 | 0.516 |
| Large | 0.564 | 0.064 | <0.001 | 0.439 | 0.688 |

\* **Adjusted mean difference in length of hospital stay**
TAVR, transcatheter aortic valve replacement; SAVR, surgical aortic valve replacement; HMO, health maintenance organization

Upon further analysis, it was observed that certain patient characteristics were associated with a shorter adjusted mean LOS. Specifically, female patients, older patients, Black and Hispanic patients, patients admitted over the weekend, Medicare patients and patients receiving early aortic valve replacements exhibited significantly shorter LOS (Table 3).

### Perioperative complications

The overall incidence of complications in the study was determined to be 19.2%. When analyzing the specific subgroups, it was found that the SAVR subgroup had an incidence of 8.4% in complications, while individuals who received TAVRs experienced a slightly lower incidence of 6.9%. Among all study participants, the most commonly observed complication was acute respiratory failure, accounting for 6.3% of cases. This was followed by the need for critical care admission at 5.9%, acute myocardial infarction at 2.6%, postoperative hemorrhage at 2.2%, and postoperative cardiac functional disturbances at 1.2%. Additionally, 3.6% of the recorded complications were categorized as unspecified. Table 4 provides a comprehensive overview of the frequency of complications observed in the study.

**Table 4.** Perioperative complications in the total study cohort: stratified by procedural approach.

| **Complications** | **Total cohort, N (%)** | **SAVR, N (%)** | **TAVR, N (%)** |
| --- | --- | --- | --- |
| Any complication | 69,820 (19.2) | 30,660 (8.4) | 25,120 (6.9) |
| Post-procedural heart failure | 729 (0.2) | 500 (0.1) | 135 (0.03) |
| Post-cardiotomy syndrome | 4,738 (1.3) | 2,270 (0.6) | 1,525 (0.4) |
| Intra-operative hemorrhage and hematoma complicating bypass or catheterization | 1,094 (0.3) | 380 (0.1) | 700 (0.2) |
| Post-operative hemorrhage | 8,055 (2.2) | 3,189 (0.9) | 4,014 (1.0) |
| Intraoperative cardiac arrest | 1,458 (0.4) | 215 (0.05) | 960 (0.3) |
| Intraoperative cardiac functional disturbances | 1,458 (0.4) | 255 (0.07) | 1,000 (0.3) |
| Postoperative cardiac functional disturbances | 4,374 (1.2) | 1,874 (0.5) | 2,225 (0.6) |
| Intraoperative cerebrovascular infarction | 145 (0.04) | 55 (0.01) | 90 (0.02) |
| Postprocedural cerebrovascular infarction | 1,823 (0.5) | 505 (0.1) | 1,285 (0.4) |
| Myocardial infarction | 9,477 (2.6) | 2,845 (0.8) | 2,560 (0.7) |
| Cardiac tamponade | 2,551 (0.7) | 1,040 (0.3) | 1,375 (0.4) |
| Infective endocarditis | 510 (0.14) | 340 (0.1) | 60 (0.02) |
| Wound dehiscence | 510 (0.14) | 420 (0.1) | 35 (<0.01) |
| Critical care admission | 21,506 (5.9) | 10,345 (2.8) | 6,745 (1.9) |
| ARDS | 364 (0.1) | 180 (0.05) | 84 (0.02) |
| Acute respiratory failure | 22,964 (6.3) | 9,240 (2.5) | 6,720 (1.8) |
| Aortic dissection | 36 (0.01) | 10 (<0.01) | 15 (<0.01) |
| Hemolytic anemia | 36 (0.01) | 20 (<0.01) | 10 (<0.01) |
| Other unspecified complications* | 13,122 (3.6) | 8,184 (2.2) | 4,360 (1.2) |

SAVR, surgical aortic valve replacement; TAVR, transcatheter aortic valve replacement; ARDS, acute respiratory distress syndrome
\*Includes intraoperative and postoperative complications not elsewhere classified

After adjustments, we discovered a significant association between SAVR and an elevated probability of various complications, including heart failure, post-cardiotomy syndrome, postoperative hemorrhage, postoperative cardiac functional disturbances, postoperative strokes, and myocardial infarction. Furthermore, SAVR was found to increase the likelihood of cardiac tamponade, infective endocarditis, critical care admission, and acute respiratory failure in comparison to medical management. Likewise, TAVR increased the probability of intraoperative cardiac arrest and functional disturbances, postprocedural strokes, postprocedural hematomas, and cardiac tamponade when compared to medical management (Table 5).

**Table 5.** Odds of perioperative complications on multivariate logistic regression analysis Complications Multivariable regression analysis.

| Complications | Multivariable regression analysis<br>aOR (95% CI; P-value) |  |
| --- | --- | --- |
|  | SAVR | TAVR |
| Any complication | 2.52 (2.29–2.78;<br>P<0.001) | 0.89 (0.80-0.97;<br>P=0.01) |
| Post-procedural heart failure | 5.42 (2.08–4.11;<br>P=0.001) | 0.75 (0.29-1.90;<br>P=0.542) |
| Post-cardiotomy syndrome | 1.75 (1.32-2.32;<br>P<0.001) | 0.69 (0.53-0.91;<br>P=0.007) |
| Intra-operative hemorrhage and hematoma<br>complicating bypass or catheterization | 1.63 (0.69-3.87;<br>P=0.268) | 1.63 (0.73-3.62;<br>P=0.232) |
| Post-operative hemorrhage | 3.87 (2.62-5.71;<br>P<0.001) | 1.15 (0.78-1.68;<br>P=0.480) |
| Accidental puncture and laceration of a circulatory<br>system organ or structure | 1.54 (0.82-2.88;<br>P=0.180) | 1.68 (0.97-2.84;<br>P=0.051) |
| Postprocedural hematoma of a circulatory system<br>organ or structure | 0.75 (0.45-1.24;<br>P=0.259) | 1.66 (1.07-2.57;<br>P=0.02) |
| Intraoperative cardiac arrest | 1.33 (1.59-2.99;<br>P=0.001) | 3.17 (1.56-6.42;<br>P<0.001) |
| Intraoperative cardiac functional disturbances | 1.32 (0.62-2.79;<br>P=0.472) | 2.76 (1.41-5.37;<br>P=0.003) |
| Postoperative cardiac functional disturbances | 2.83 (1.83-4.37;<br>P<0.001) | 2.13 (1.39-3.27;<br>P=0.001) |
| Intraoperative cerebrovascular infarction | 4.66 (0.86-25.22;<br>P=0.007) | 2.63 (0.57-12.10;<br>P=0.215) |
| Postprocedural cerebrovascular infarction | 4.23 (2.11-8.51;<br>P<0.001) | 3.36 (1.78-6.35;<br>P<0.001) |
| Myocardial infarction | 2.10 (1.78-2.48;<br>P<0.001) | 0.98 (0.84-1.15;<br>P=0.810) |
| Cardiac tamponade | 2.41 (1.32-1.89;<br>P=0.004) | 1.79 (1.04-3.07;<br>P=0.030) |
| Infective endocarditis | 2.82 (1.34-5.92;<br>P=0.01) | 0.62 (0.25-1.53;<br>P=0.301) |
| Wound dehiscence | 1.70 (0.65-4.43;<br>P=0.277) | 0.16 (0.05-0.47;<br>P=0.001) |
| Critical care admission | 1.40 (1.20-1.64;<br>P<0.001) | 0.54 (0.46-0.63;<br>P<0.001) |
| ARDS | 0.68 (0.24-1.91;<br>P=0.463) | 0.41 (0.13-1.32;<br>P=0.137) |
| Acute respiratory failure | 1.40 (1.23-1.60;<br>P<0.001) | 0.56 (0.48-0.64;<br>P<0.001) |
| Other unspecified complications | 8.03 (6.25-10.30;<br>P<0.001) | 1.93 (1.51-2.48;<br>P<0.001) |

SAVR, surgical aortic valve replacement; TAVR, transcatheter aortic valve replacement; aOR, adjusted odds ratio

In a comparative analysis, SAVRs exhibited a higher likelihood of postoperative arrhythmias, acute respiratory failures, postoperative strokes, and cardiac tamponades when contrasted with TAVRs. Conversely, TAVRs were associated with a greater likelihood of postoperative hematomas, intraoperative arrhythmias, and cardiac arrests relative to SAVRs. In comparison to SAVRs, TAVRs demonstrate a lower probability of post-cardiotomy syndromes, wound dehiscence, critical care admission, and acute respiratory failure.

Overall, TAVRs exhibited a lower likelihood of experiencing any complication when compared to medical management or SAVRs.

### Total hospital charges

The overall mean hospital charge for aortic stenosis patients admitted during the study period was $186,494 ± $1,908. Patients managed solely with medical therapy spent less in hospital costs compared to patients receiving aortic valve replacements ($86,192 ± $1,249 vs. $211,431 ± $2,203).

For TAVR, the average hospital cost was $209,513 ± $2,713 while for SAVR it was $215,366 ± $2,183. Patients who had aortic valve replacement within 3 days of admission paid significantly less in hospital costs compared to those who had their surgery at a later time ($144,469 vs. $202,497).

After controlling for factors such as procedural approach, complications, prolonged hospital stay (≥ 18 days), delayed surgery (≥ 3 days from admission), early surgery (within 3 days of admission), and various other patient and hospital-level variables, it was observed that transcatheter aortic valve replacement (TAVR) was associated with a higher average total hospital cost in comparison to surgical aortic valve replacement (SAVR) ($161,139 ± $4,381 vs $148,836 ± $4,011; P<0.001). Notably, several factors were identified as contributors to the increased hospital costs in the study. These factors included prolonged hospital stay, the occurrence of any complication during the admission, admission to medium or large teaching hospitals, admission to hospitals located in the western region, belonging to a median annual income quartile of ≥ $74,000, being of Hispanic race, and having a higher Charlson comorbidity index.

## DISCUSSION

The index study aimed to compare the outcomes and care costs between SAVR and TAVR for AS hospitalizations. Our findings revealed several key insights into the sociodemographic characteristics, inpatient mortality rates, LOS, perioperative complications, and total hospital charges associated with these interventions.

The sociodemographic characteristics of the study population provided valuable context for understanding the patient profiles. The majority of admissions were in older individuals, with a mean age of 76 years, consistent with the current evidence of AS predominantly affecting older individuals.^19,20^ Interestingly, there were distinct age distributions between the SAVR and TAVR subgroups, with SAVR patients tending to be younger (mean age of 69 years) compared to TAVR patients (mean age of 79 years). This age difference is likely reflective of the evolving guidelines and eligibility criteria for these procedures, with TAVR traditionally being reserved for older patients with high or prohibitive risk for SAVR.^21–23^ Additionally, a higher proportion of men were observed in the study cohort, consistent with AS being more prevalent in males. At a cellular level, it has been reported that males with AS exhibit an upregulation of collagen and metalloproteinase genes, and comparatively, male hearts with AS demonstrate a higher presence of fibrosis, larger left ventricular volumes, and higher wall tension when compared to females.^24,25^ Racially, similar to existing literature, the majority of patients were White Americans.^26^ This highlights a critical concern about equity in healthcare access and the need for diverse representation in future studies to assess potential disparities in treatment and outcomes.

Our study demonstrated differences in inpatient mortality rates among the treatment groups. The overall mortality rate in our study cohort was 2%, with SAVR and TAVR accounting for 28.6% and 29.9% of all mortalities, respectively. Adjusted analysis revealed that TAVR was associated with significantly lower odds of in-hospital mortality compared to medical management, whereas SAVR did not show a significant difference. These findings suggest that TAVR may confer a survival advantage over medical management, aligning with previous studies that have shown favorable outcomes with TAVR in high-risk and older patients.^27^ The lack of significant difference in mortality between SAVR and medical management in this study may be attributed to several factors, including patient selection and the influence of other procedure-related variables not fully accounted for in our analysis.

The LOS is an important indicator of healthcare resource utilization and patient recovery. Our results demonstrated that patients undergoing SAVR had a longer average length of stay (8.1 days) compared to those undergoing TAVR (3.5 days). Furthermore, both SAVR and TAVR patients had longer lengths of stay compared to patients managed medically. After adjusting for confounding factors, TAVR was associated with a significantly shorter LOS compared to SAVR. These findings are consistent with previous studies suggesting that TAVR may enable faster recovery and shorter hospital stays compared to SAVR, likely due to the less invasive nature of the procedure.^28^ However, there is evidence in the literature that certain procedural factors may modify the effect of TAVR on LOS. For example, conscious sedation has been associated with shorter LOS while a transapical approach has been associated with prolonged LOS.^29^ Additionally, we identified several patient characteristics associated with shorter lengths of stay, such as female sex, older age, and certain racial and insurance factors, warranting further investigation to understand the underlying mechanisms.

Perioperative complications are critical indicators in evaluating the safety and effectiveness of interventions. Our study revealed an overall incidence of complications of 19.2% in the study cohort. SAVR patients had a higher incidence of complications (8.4%) compared to TAVR patients (6.9%). The adjusted analysis demonstrated that both SAVR and TAVR procedures were associated with specific complications, with SAVR patients being at higher risk for certain complications, including heart failure, post-cardiotomy syndrome, and cardiac tamponade. Conversely, TAVR patients had a higher likelihood of intraoperative arrhythmias, post-procedural hematomas, and functional disturbances. TAVR demonstrated a lower likelihood of experiencing any complication when compared to both SAVR and medical management, suggesting that TAVR may offer a safer alternative in high-risk patient populations.

Total hospital charges play a crucial role in healthcare cost assessments. Our study revealed that patients managed medically had lower total hospital costs compared to those who received aortic valve replacements. Furthermore, TAVR patients had higher average total hospital costs compared to SAVR patients. After adjusting for relevant factors, TAVR was associated with significantly higher average total hospital costs compared to SAVR. Several factors may have contributed to the increased hospital costs, including the occurrence and implications of managing specific complications, specific hospital characteristics, higher comorbidity burdens, the cost of deploying TAVR technology, the cost of TAVR valves, and income disparities.

Despite higher initial procedural and index hospitalization costs, recent evidence suggests that TAVRs may be more cost-effective in the long term compared to SAVRs. A recent U.S. PARTNER 3 trial enrolled 979 AS patients to compare TAVR with the SAPIEN 3 valve to SAVR using any bioprosthetic valve. Index hospitalization costs were slightly higher for TAVR, mainly due to procedural expenses, but overall hospitalization costs were lower for TAVR patients due to shorter hospital stays, especially in the ICU. At the 2-year follow-up, TAVR showed a trend towards lower costs compared to SAVR. Particularly, patients with severe symptoms and lower quality of life scores were found to benefit more in terms of cost savings with TAVR.^30^ These findings highlight the economic implications associated with the different treatment approaches and emphasize the need for further cost-effectiveness analyses to guide decision-making in the management of aortic stenosis.

In interpreting our findings, it is essential to acknowledge certain limitations. Our study relied on retrospective data from the study period, which may be subject to inherent biases and limitations associated with administrative database analyses such as limited data on disease severity. Additionally, owing to the nature of the data source, there may be unmeasured intraoperative variables that could influence the observed associations. Furthermore, our study focused on in-hospital outcomes, limiting our ability to assess post-discharge outcomes and long-term survival rates. Despite these limitations, we made meticulous efforts to minimize the effects of residual confounding through the use of rigorous statistical methods and large sample size.

## CONCLUSION

In conclusion, our comparative analysis of outcomes and care costs between SAVR and TAVR for AS hospitalizations yielded valuable insights. TAVR demonstrated a significant reduction in the odds of in-hospital mortality compared to medical management, whereas SAVR did not show a significant difference. TAVR was associated with shorter adjusted lengths of stay and a lower likelihood of experiencing complications compared to both SAVR and medical management. However, TAVR was associated with higher total hospital costs compared to SAVR or medical management. These findings underscore the importance of considering individual patient characteristics, procedural factors, and cost implications when making treatment decisions for patients with aortic stenosis. Future studies with long-term follow-up data are warranted to provide a comprehensive understanding of post-discharge outcomes and long-term cost-effectiveness in this patient population.

## Data Availability

In compliance with the copyright restrictions set by the AHRQ pertaining to the distribution of HCUP databases, the database utilized in this research will not be made publicly available by the authors. However, all NIS datasets are publicly available through the authorized HCUP central distributor upon request at https://hcup-us.ahrq.gov/tech_assist/centdist.jsp. or via direct email to the AHRQ at

## Funding Information

This study did not receive any funding

## Informed consent

The absence of patient and hospital-level identifiers in the NIS precludes obtaining informed consent. Nevertheless, this study diligently adheres to the standards mandated by the Agency for Healthcare Research and Quality (AHRQ), which regulates the use of the Nationwide Inpatient Sample.

## Research involving human participants, their data, or biological material

This research did not involve any human subjects and no biological material was collected at any stage of the study.

## Notes

**Conflict of interest:** The authors report no conflicting interests

### Competing Interest Statement

The authors have declared no competing interest.

### Author Declarations

The study used ONLY publicly available NIS data that were originally available at: https://hcup-us.ahrq.gov/tech_assist/centdist.jsp.

